# Preserving a decade of machine-learning validity across a ground-up refactoring: a five-tier automated validation of RCT-Reviewer, an independent modernization of RobotReviewer

**DOI:** 10.64898/2026.09.08.26362546

**Authors:** Vihaan Sahu, Ajit Sahu

## Abstract

Evidence-synthesis teams increasingly depend on machine-learning tools to automate risk-of-bias assessment, but these tools frequently rely on unrunnable, deprecated software stacks. Refactoring legacy tools into modern environments is essential for maintenance, yet introduces a critical risk: silently invalidating published performance metrics. Without rigorous validation, systematic reviewers cannot trust that modernized tools retain their predecessors’ behavior. We present a five-tier validation framework applied to RCT-Reviewer, an independent ground-up modernization of the widely used RobotReviewer system. To prove preservation, we engineered a compatibility shim to execute the original 2017 code alongside the modernized tool using byte-identical weight files. The framework evaluated predictive validity on a human-labelled benchmark, quantified the cost of dropping obsolete ensemble components, proved bit-exact fidelity, tested parser robustness at scale, and included an external validation arm evaluating agreement on 313 trials against a recent reference standard. A control experiment showing 100% agreement between legacy and modernized implementations on identical text proved that variance versus published values is driven by PDF provenance (open-access versus publisher typeset) rather than codebase refactoring. The modernized tool perfectly reproduced original risk-of-bias judgements in 6,018 comparisons. While the screening classifier experienced a bounded performance reduction (F1 0.969 to 0.925) from removing obsolete components, the model’s core mathematical validity is preserved. These results establish that reviewers can deploy a maintainable modernization with a clearly bounded performance trade-off, offering a reusable validation framework for clinical NLP modernizations. Future work will explore retraining obsolete neural network components to close the performance gap.

## 1. Introduction

Systematic reviews underpin evidence-based medicine, but their creation is bottlenecked by labor-intensive tasks like risk-of-bias (RoB) assessment.[1,14] Machine learning (ML) tools, such as RobotReviewer, were developed to automate RoB extraction and have been validated in real-world review pipelines.[2,3] However, the original tool relies on deprecated infrastructure (Python 3.6, TensorFlow 1.x, and Java-based GROBID parsing). As software stacks evolve, these tools face “software rot,” becoming unrunnable and threatening the reproducibility of evidence-synthesis workflows.

Refactoring legacy ML tools into modern environments (e.g., Python 3.12, native PDF parsing) is essential for maintenance, but introduces a critical methodological risk: silently invalidating the published performance metrics. A refactored tool might parse text differently, drop unsupported ensemble components, or alter tokenization, thereby changing clinical judgements. Previous evaluations of RobotReviewer, including those by Gates et al. (2018), Armijo-Olivo et al. (2020), Hirt et al. (2021), and the largest validation to date by Tian et al. (2024), established the tool’s baseline reliability against human reviewers.[5–8] However, none of these studies evaluated whether this validated behavior could survive a ground-up software refactoring, nor did they control for how PDF provenance affects RoB judgements.

This manuscript demonstrates that a decade of machine-learning validity can be preserved across a ground-up software refactoring. To address the unrunnable state of the original tool, the lead author (VS) independently developed RCT-Reviewer, a GPL-3.0 modernization of RobotReviewer.[15] To prove validity, we developed a five-tier automated validation framework. The framework evaluates predictive validity against a benchmark, quantifies the cost of dropping dead ensemble components, proves bit-exact fidelity against the original 2017 code, tests robustness at scale, and includes an external, human-referenced validation tier using the dataset published recently in this journal by Tian et al.[8]

The framework is the methodological contribution; the tool is the demonstration. By proving that mathematical fidelity is maintainable by weight-identity, we isolate PDF provenance as a hidden variable in automated RoB evaluation and establish a reusable standard for validating clinical NLP modernizations.

## 2. Methods

We evaluated RCT-Reviewer against the original RobotReviewer (2017 commit) using a dedicated, reproducible validation harness (https://github.com/RCT-Reviewer/Validation). All code is seeded and hash-verified. While it is theoretically possible to execute the original tool in an isolated legacy environment (e.g., a Python 3.6 container), doing so relies on end-of-life, insecure dependencies and defeats the purpose of software modernization. Furthermore, running the tools in separate environments would introduce environment-level variance. To achieve a true apples-to-apples comparison, we engineered a compatibility shim (validation_shim.py) to run the legacy 2017 code directly inside the modern Python 3.12 environment without altering its numerics. This shim applies non-numerical patches (e.g., Keras stubs, scikit-learn kwarg translation) and redirects the legacy code to load the exact same byte-identical weight files as the modern tool, ensuring that any differences in output are attributable to the tool’s logic rather than diverging dependencies.

The validation consists of five tiers:

- **Tier A (Predictive Validity):** We scored the 751-record Clinical Hedges MEDLINE benchmark through the refactored classifier and compared results to human labels across five metrics (sensitivity, specificity, F1, κ, AUC). We also compared the scores to the executed original code using a maximum absolute delta (max |Δ|) acceptance gate to ensure numerical fidelity.
- **Tier B (CNN Ablation):** The original tool used an SVM+CNN+publication-type (ptyp) ensemble.[4] Because the CNN relies on dead TensorFlow 1.x, it cannot be executed in modern maintained environments. We compared the refactored SVM-only decisions against the stored decisions of the full ensemble (originally generated by the tool’s developers and deposited alongside the 2017 codebase) using McNemar’s exact test to quantify the cost of ablation.
- **Tier C (RoB Fidelity):** Because the original CNN relies on dead TensorFlow 1.x, we used the compatibility shim to run the 2017 pipeline in an SVM-only capacity, creating an apples-to-apples comparison against the refactored SVM-only tool. We ran both pipelines over 1,003 documents × 6 RoB domains (6,018 comparisons) and measured judgement agreement, top-3 evidence-sentence Jaccard, and sentence decision score deltas.
- **Tier D (Parser Robustness):** We parsed 1,000 open-access PDFs (12,060 pages) from Europe PMC, a corpus size consistent with recent automated systematic review tool evaluations.[12] We measured parse success rate, time, and linear scaling.
- **Tier E (External Human-Referenced Validation and Control Experiment)**: Tian et al. evaluated publisher PDFs. Because our harness can only legitimately retrieve open-access PMC text, a gap versus published values is expected from the input difference alone. To prove the gap is the PDF source and not the refactoring, we re-ran the original 2017 code (via the shim) on the identical PMC text that RCT-Reviewer judged, isolating PDF provenance as the only variable.

## 3. Results

### Tier E: External Human-Referenced Validation

On the 313-trial open-access subset, RCT-Reviewer agreed with the human consensus at κ = 0.26, 0.20, 0.48, and 0.12 across the four RoB domains (concordance 60–76%). This is within the range Tian et al. published for the original tool on publisher PDFs (κ = 0.25–0.59, concordance 63–83%), and aligns with broader evaluations of RoB automation which note moderate agreement depending on the domain.[7,10]

The control phase confirmed the mechanism: the original 2017 implementation (run in SVM-only mode via the shim), when executed on the identical PMC text, agreed with RCT-Reviewer in 100.0% of domain judgements and showed identical human κ. External fidelity versus the original’s deposited publisher-PDF labels was 78.9%. This proves the difference versus published values is attributable to the PDF source (open-access versions vs publisher PDFs), not the refactoring. The per-domain agreement is detailed in Table 1.

(Figure 1)

**Figure 1:**
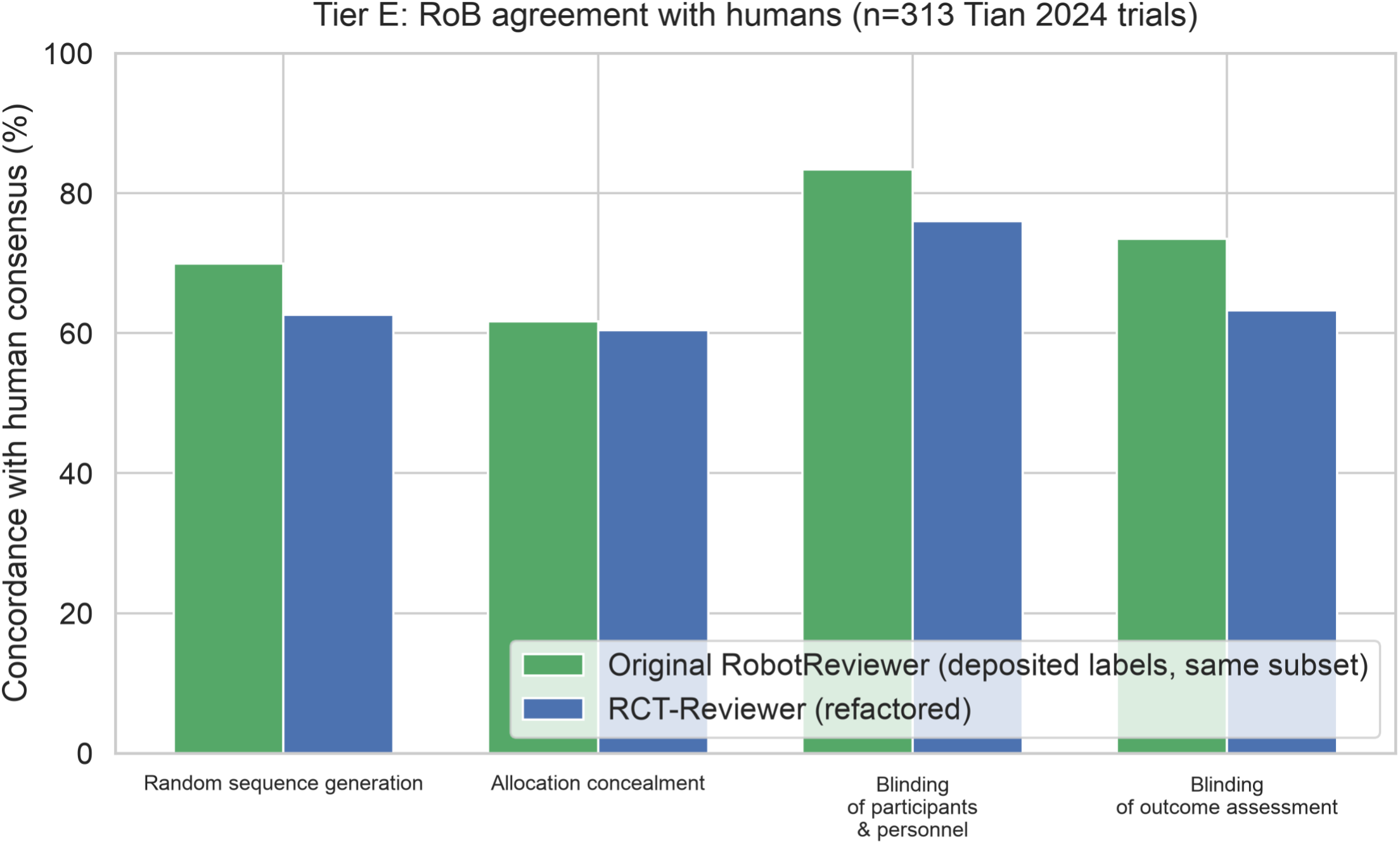
Per-domain concordance with the human consensus on the Tian 2024 open-access subset (n=313). RCT-Reviewer (blue) vs the original RobotReviewer’s deposited labels (green) on the identical trials. The control phase additionally showed the two implementations agree on 100% of judgements when given the same text. *(File: figure_tier_e_human_concordance.png)*

**Table 1:** Tier E - Per-Domain Agreement with Human Consensus (Tian 2024 Subset, n=313)

| DOMAIN | RCT-REV VS HUMAN: K (95% CI) | CONCORDANCE (95% CI) | ORIGINAL RR (DEPOSITED LABELS, PUBLISHER PDFS) VS HUMAN: K | TIAN PUBLISHED K (FULL N=1955) |
| --- | --- | --- | --- | --- |
| Random sequence | 0.26 (0.15–0.36) | 62.6 (57.1–67.8) | 0.40 | 0.46 |

| **DOMAIN** | **RCT-REV VS**<br>**HUMAN: K**<br>**(95% CI)** | **CONCORDANCE**<br>**(95% CI)** | **ORIGINAL RR**<br>**(DEPOSITED**<br>**LABELS,**<br>**PUBLISHER**<br>**PDFS) VS**<br>**HUMAN: K** | **TIAN**<br>**PUBLISHED K**<br>**(FULL N=1955)** |
| --- | --- | --- | --- | --- |
| generation |  |  |  |  |
| Allocation<br>concealment | 0.20 (0.10–0.30) | 60.4 (54.9–65.6) | 0.25 | 0.25 |
| Blinding of<br>participants/pers<br>onnel | 0.48 (0.38–0.57) | 76.0 (71.0–80.4) | 0.58 | 0.59 |
| Blinding of<br>outcome<br>assessment | 0.12 (0.01–0.23) | 63.3 (57.8–68.4) | 0.31 | 0.27 |

### Tier C: Mathematical Fidelity

The refactored RoB pipeline perfectly reproduced the SVM-only judgements of the original 2017 code in 6,018/6,018 document × domain comparisons (κ = 1.0). Sentence scores were identical, and vectorizer matrices were byte-identical (14/14 probes). All four model weight files were verified byte-identical via SHA-256 hashing against the original Git LFS object hashes. Published RoB accuracy therefore transfers by weight identity, meaning the mathematical behavior of the 2017 model is perfectly preserved in the modern environment. The exact concordance and fidelity metrics are presented in Table 2.

**Table 2:** Tier C - Risk-of-Bias Pipeline Fidelity (Original vs. Refactored)

| **COMPARISON / METRIC** | **VALUE** |
| --- | --- |
| Total comparisons (documents × 6 domains) | 6,018 |
| Judgement agreement (95% CI) | 100.0 (99.9–100.0) |
| Cohen's kappa | 1.0000 |
| Max $\vert \Delta$ sentence score\| | 0.000e+00 |
| Vectorizer matrices identical | True (14 of 14 probes) |
| Model weight files verified (SHA-256) | 4/4 byte-identical |

### Tier A: Predictive Validity

On the human-labelled Clinical Hedges benchmark (n=751), the refactored tool achieved strong predictive validity, consistent with prior automated screening tools.[9] The tool achieved a sensitivity of 94.1% (91.4– 96.0), specificity of 88.1% (84.2–91.2), F1 of 0.925, Cohen’s κ of 0.826, and ROC AUC of 0.966. Fidelity versus the executed original code was exact (max |Δ| = 0.0, agreement 100%), confirming that the removal of dead dependencies did not alter the classifier’s decision boundaries. Performance metrics are detailed in Table 3.

(Figure 2 and Figure 3)

**Figure 2:**
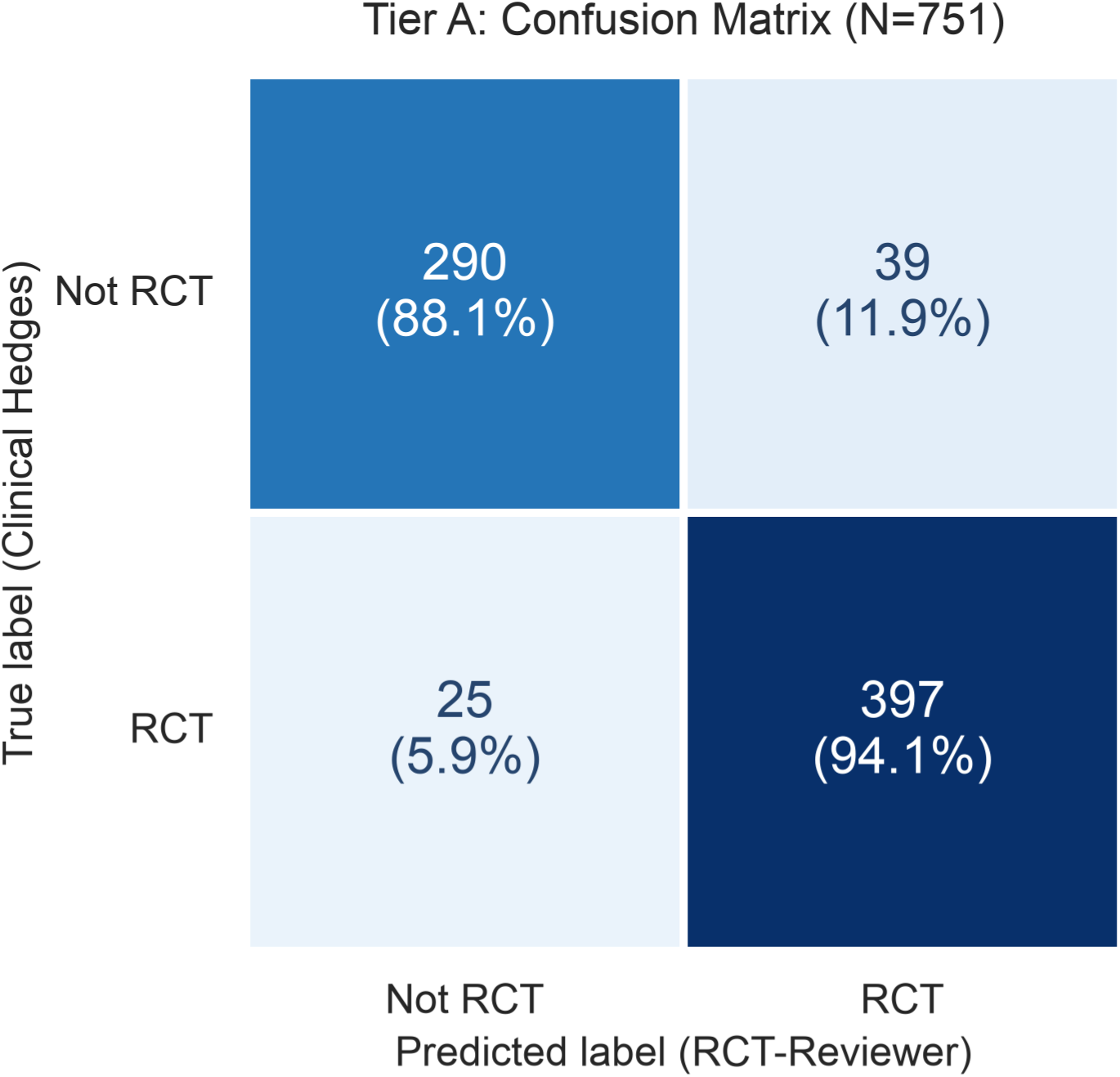
Confusion matrix of the refactored SVM classifier against human labels (Clinical Hedges, n=751). Diagonal cells are correct calls (290 not-RCTs rejected, 397 RCTs accepted); off-diagonal cells are errors (39 wrongly retained, 25 missed). Cell percentages represent specificity (88.1%, top row) and sensitivity (94.1%, bottom row). *(File: figure_tier_a_confusion_matrix.png)*

**Figure 3:**
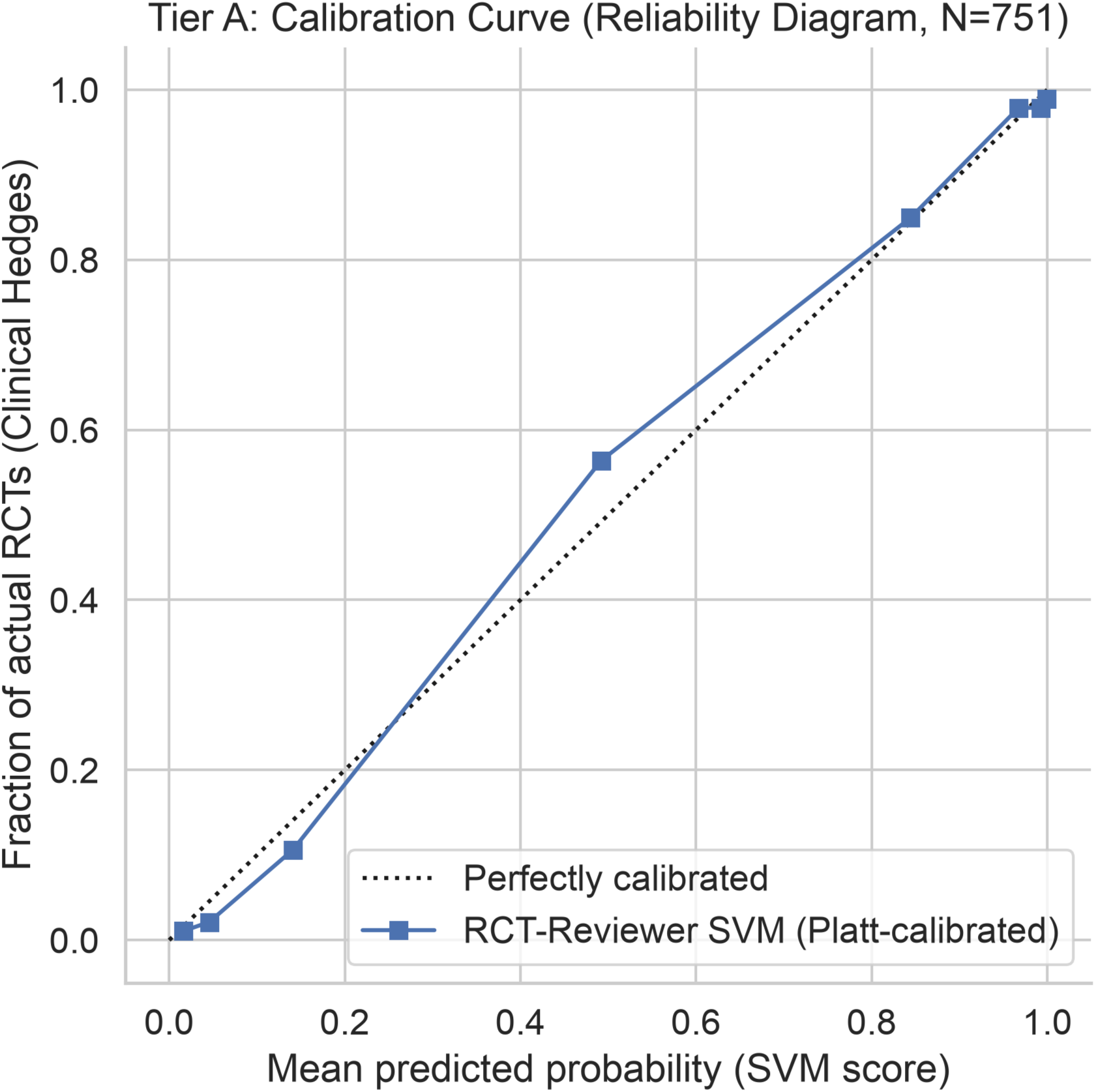
Reliability diagram of the SVM score after in-sample Platt scaling. Points near the diagonal indicate the calibrated score reads as a probability. *(File: figure_tier_a_calibration.png)*

**Table 3:** Tier A - RCT Classifier Performance on Clinical Hedges Benchmark (n=751)

| METRIC | VALUE (95% CI) |
| --- | --- |
| Sensitivity | 94.1 (91.4–96.0) |
| Specificity | 88.1 (84.2–91.2) |
| Accuracy | 91.5 (89.3–93.3) |
| PPV | 91.1 (88.0–93.4) |
| NPV | 92.1 (88.5–94.6) |
| F1 Score | 0.925 (0.907–0.943) |
| Cohen's Kappa | 0.826 (0.786–0.866) |
| ROC AUC | 0.9658 |

### Tier B: SVM/CNN Ablation

The SVM-only pipeline achieved 92.5% decision agreement with the original full SVM+CNN+ptyp ensemble (F1 0.925 vs 0.969; McNemar p < 0.0001). Three-arm attribution showed the SVM-only agrees 94.5% with SVM+CNN (no ptyp), indicating that publication-type features (not the CNN) drive most of the performance gap. Both dropped components are unrunnable in a maintained environment. This quantifies the trade-off of modernization: a measurable but statistically bounded decrease in ensemble performance in exchange for full software maintainability. The ablation results are summarized in Table 4.

(Figure 4)

**Figure 4:**
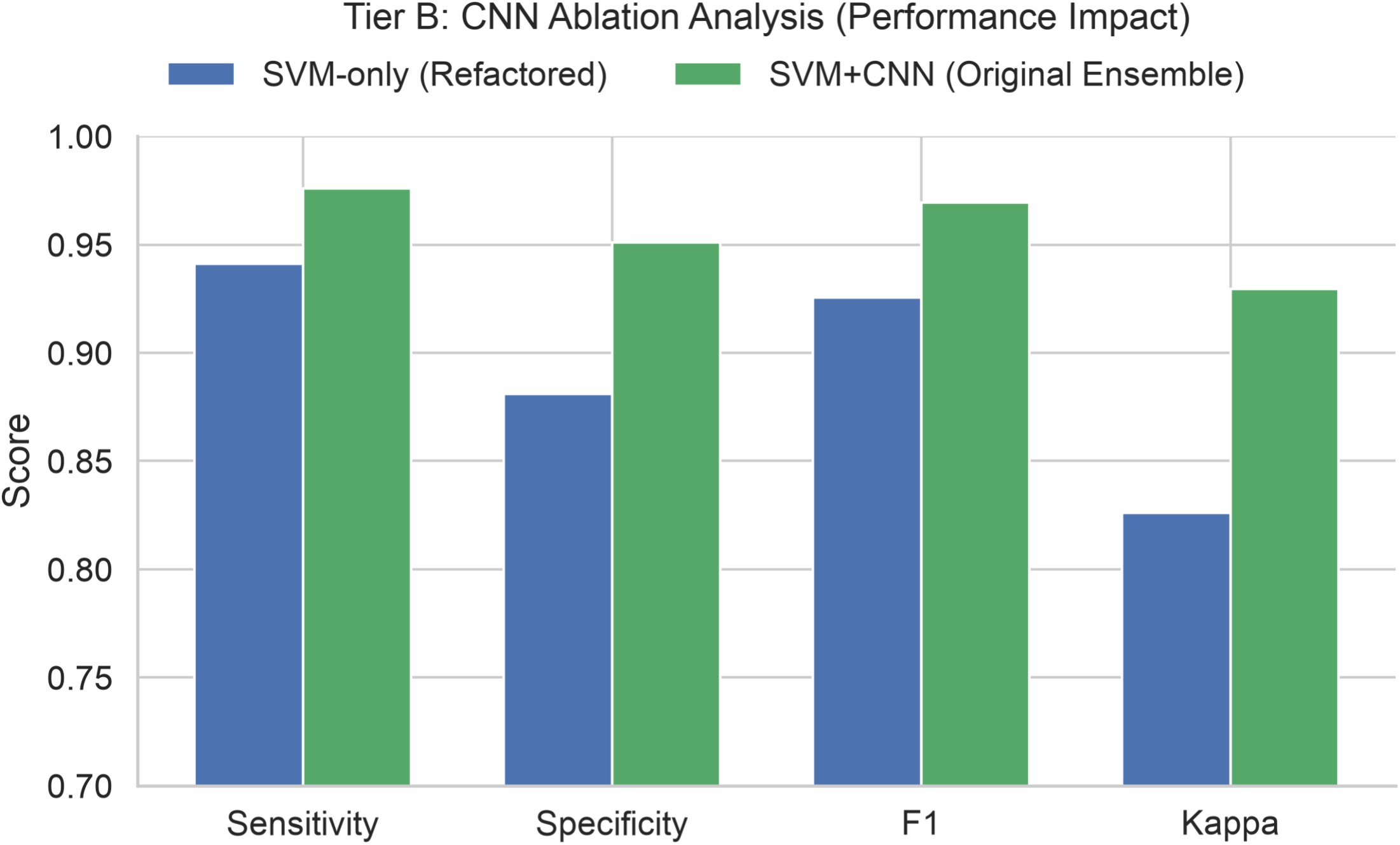
CNN ablation comparison showing the measurable cost of removing the unmaintainable TensorFlow CNN. SVM-only (refactored) vs the original full SVM+CNN ensemble, side by side. F1 0.925 vs 0.969 (McNemar p < 0.0001); most of the gap comes from the publication-type features. *(File: figure_tier_b_ablation.png)*

**Table 4:** Tier B - CNN Ablation (SVM-only vs. SVM+CNN Ensemble)

| MODEL CONFIGURATION | SENSITIVITY<br>(95% CI) | SPECIFICITY<br>(95% CI) | F1 SCORE<br>(95% CI) |
| --- | --- | --- | --- |
| SVM-only (Refactored) | 94.1 (91.4–96.0) | 88.1 (84.2–91.2) | 0.925 (0.907–0.943) |
| SVM+CNN (no ptyp) | 95.5 (93.1–97.1) | 94.5 (91.5–96.5) | 0.956 (0.942–0.969) |
| Full Ensemble (SVM+CNN+ptyp) | 97.6 (95.7–98.7) | 95.1 (92.2–97.0) | 0.969 (0.957–0.980) |
| *Note: McNemar's Test (SVM vs Full Ensemble) p-value < 0.0001.* |  |  |  |

### Tier D: Parser Robustness

All 1,000/1,000 PDFs parsed successfully (95% CI 99.6–100.0) using the new PyMuPDF-based parser, which replaces the legacy Java/GROBID dependency. Median processing time was 1.66 seconds per PDF (IQR 1.32– 2.06), with processing time scaling linearly with document length (Pearson r = 0.93) and no blow-ups on the longest documents (max 7.0 s). The lexical plausibility of extracted evidence sentences across RoB domains is shown in Table 5, reflecting how often modern open-access papers explicitly describe these methodological domains.

(Figure 5 and Figure 6)

**Figure 5:**
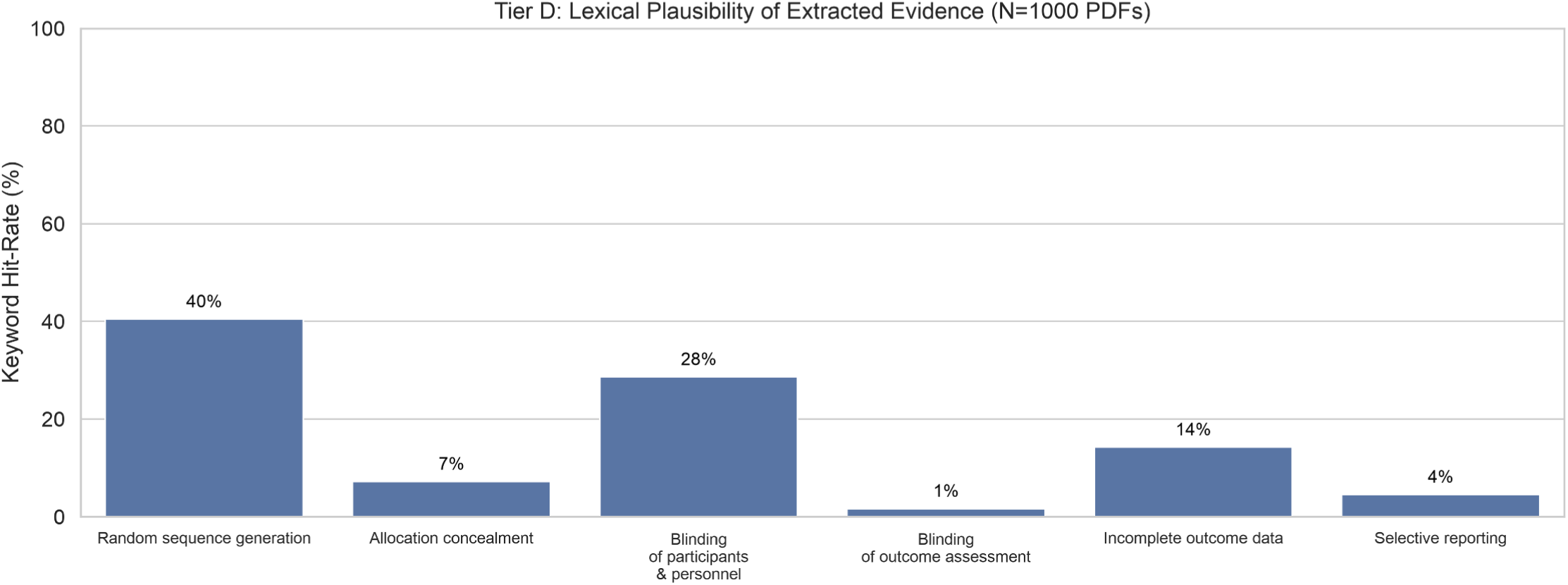
Lexical plausibility check of extracted evidence. Keyword hit-rate of the top-3 highlighted evidence sentences per RoB domain (3,000 sentences per domain). Low rates reflect how rarely papers describe those domains explicitly. *(File: figure_tier_d_keywords.png)*

**Figure 6:**
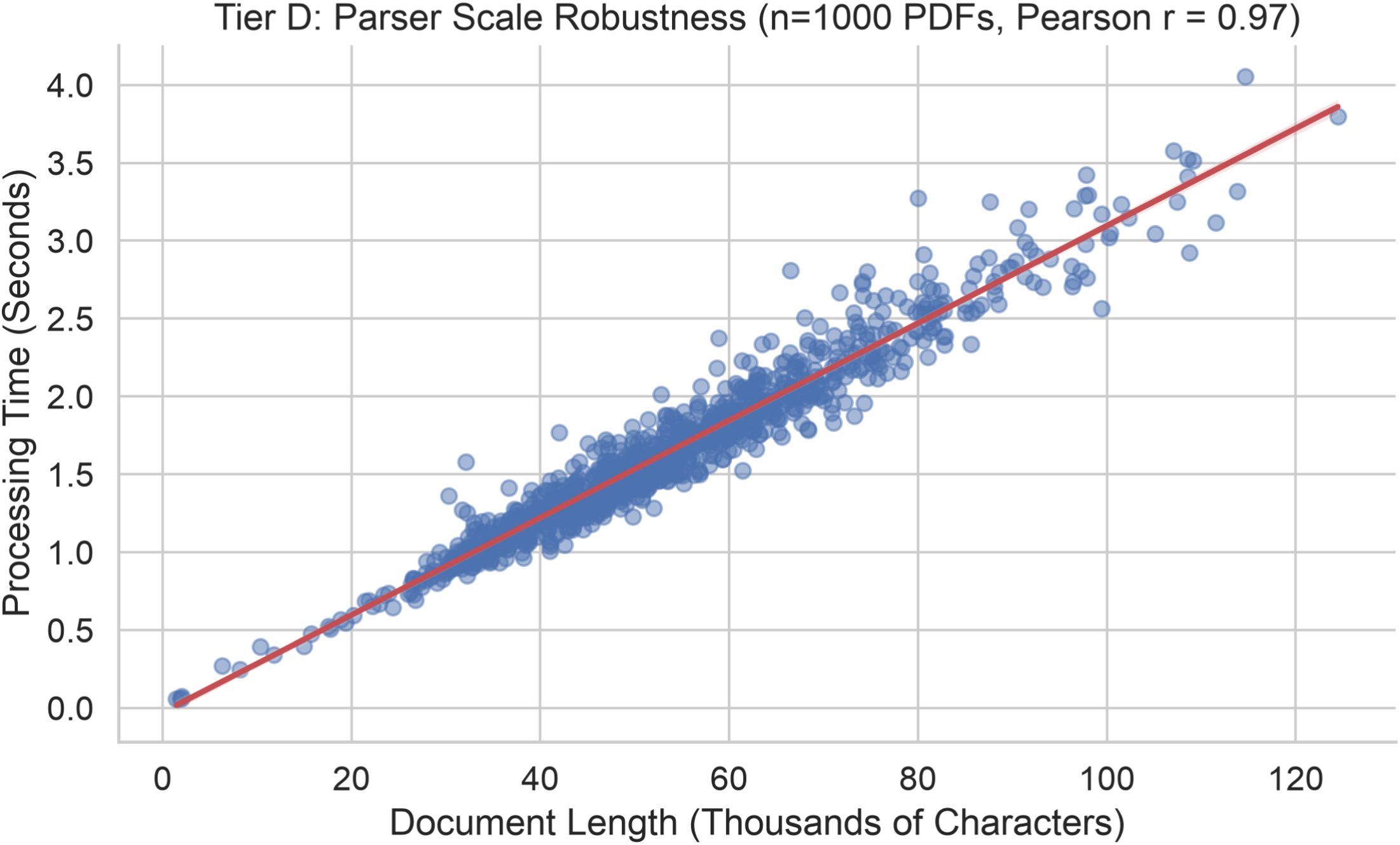
Parse time vs document length across all 1,000 PDFs. Linear scaling (Pearson r = 0.93) with no blow-ups (max 7.0 s). *(File: figure_tier_d_scale.png)*

**Table 5:** Tier D - Lexical Plausibility of Extracted Evidence (1,000 PDFs)

| RISK OF BIAS DOMAIN | KEYWORD HIT-RATE | SNIPPETS EVALUATED |
| --- | --- | --- |
| Random sequence generation | 40.5% | 3,000 |
| Allocation concealment | 7.2% | 3,000 |
| Blinding of participants and personnel | 28.6% | 3,000 |
| Blinding of outcome assessment | 1.6% | 3,000 |
| Incomplete outcome data | 14.2% | 3,000 |
| Selective reporting | 4.5% | 3,000 |

## 4. Discussion

We demonstrate that a decade of machine-learning validity can be preserved across a ground-up software refactoring. By proving bit-exact fidelity (Tier C) and weight identity, we establish that published accuracy metrics transfer without needing to re-collect expensive ground-truth RoB labels. This is a critical achievement in the current software landscape, where many evidence-synthesis tools suffer from “software rot” and lack feature completeness, leaving reviewers with fewer maintained options.[12] The RCT-Reviewer tool, a modernized standalone implementation, ensures that the evidence-synthesis community retains access to a validated, GPL-3.0 licensed resource without relying on deprecated Java or TensorFlow environments.[15]

The most significant methodological finding for the evidence-synthesis community is the role of PDF provenance. Automated RoB evaluations that draw text from open-access repositories (like PMC) will inherently display lower concordance with human reviewers than evaluations using the publisher’s typeset PDFs, regardless of the ML implementation. This control experiment isolates the supply chain as a variable and suggests that review teams deploying automated RoB tools must standardize their PDF sources. As previous real-world implementations of RobotReviewer have shown, the utility of automated tools is highly dependent on input quality and formatting.[13] Furthermore, while classifier reliability can vary significantly across medical domains[11] and reviewer trust in automation remains a known barrier to adoption,[10] proving mathematical fidelity to validated artifacts ensures that remaining trust gaps are attributable to data inputs, not the underlying algorithm.

While this case study focuses on RobotReviewer and the Cochrane Risk of Bias (RoB 1) tool,[14] the validation framework itself is generalizable to any evidence-synthesis ML tool facing software rot. For example, if a legacy RoB 2 classifier[16] is modernized, Tier A would benchmark its baseline screening accuracy, Tier B would quantify any loss from dropping obsolete components, Tier C would prove bit-exact fidelity via weight-hashing, Tier D would test parser robustness at scale, and Tier E would isolate whether disagreements with human reviewers are due to the codebase or the PDF supply chain. By isolating code refactoring from data source variance, this methodology ensures that software modernization does not silently corrupt clinical judgements.

## Limitations

The 1,000-PDF Tier D corpus is self-selected by the tool’s own SVM (dogfooding) and includes some trial protocols. Tier D measures parser success rate and speed, but does not formally benchmark extraction accuracy against the original GROBID parser across the full corpus. However, an n=1 GROBID comparison documents a genuine PyMuPDF text-extraction gap, indicating that formal text-extraction accuracy comparisons should be a focus of future tool development. The Platt calibration is fit in-sample (optimistic). No new RoB ground truth was collected; RoB validity transfers by weight identity rather than being re-measured against new human labels. Tier E is bounded to 313 trials due to the open-access availability constraints of the Tian dataset. Because this subset is restricted to open-access trials, it may not be perfectly representative of the full RCT population (e.g., potential biases toward more recent or larger trials). However, this sample size aligns with established bounded evaluations (e.g., Hirt et al. n=190; Armijo-Olivo et al. N=393), and the primary goal of Tier E was to isolate PDF provenance, not to establish a new clinical baseline.

## Future Directions

While this work successfully preserves the validated SVM core of the original tool, the measurable cost of dropping the dead CNN ensemble component (Tier B) presents an avenue for enhancement. The authors plan to investigate methods to further improve the tool’s precision, including seeking access to the exact paper data used to train the original CNN, with the aim of retraining the network within the modern environment. Additionally, exploring contemporary NLP architectures may yield a more accurate and refined tool that closes the remaining performance gap while maintaining full software maintainability.

## 5. Conclusion

The refactored tool is mathematically the original. The five-tier validation framework, culminating in a PDF-provenance control experiment, provides a reusable template for ensuring that software modernization does not corrupt evidence-synthesis pipelines. By proving bit-exact fidelity and isolating PDF provenance as a hidden variable in automated risk-of-bias assessment, we establish a rigorous standard for validating clinical NLP modernizations. The modernized, GPL-3.0 tool is freely available and requires no Java or legacy TensorFlow dependencies, ensuring long-term maintainability for evidence-synthesis teams.

## Supplementary Materials

The following supporting information is available as supplementary files attached to this submission:

- **master_summary.csv:** Headline numbers for all validation tiers.
- **provenance.json:** Run timestamp, package versions, and SHA-256 hashes of every model weight file.
- **tier_ab_records.csv & tier_ab_summary.json:** Per-record scores and summary statistics for Tiers A and B (Clinical Hedges benchmark, n=751).
- **tier_c_domain_comparisons.csv & tier_c_summary.json:** Per-document × domain comparisons for Tier C (6,018 comparisons).
- **tier_d_documents.csv, tier_d_keyword_hits.csv, & tier_d_summary.json:** Parser robustness metrics and lexical plausibility checks for the 1,000-PDF corpus.
- **tian_agreement.csv, tian_rr_judgments.csv, tian_resolution.csv, tian_control.csv:** Per-record agreement, tool judgments, citation resolutions, and control experiment data for the Tier E external human-referenced validation (n=313).

## Ethics

Not applicable. This study involves no human or animal subjects; all data were derived from publicly available published literature and open-source software repositories.

## CRediT Taxonomy

Vihaan Sahu: Conceptualization, Methodology, Software, Validation, Formal Analysis, Investigation, Data Curation, Writing - Original Draft, Visualization. Ajit Sahu: Methodology, Validation, Writing - Review & Editing. The manuscript was reviewed and finalized by both authors.

## Use of Artificial Intelligence (AI) Tools

The authors disclose the use of artificial intelligence (AI) tools, particularly ChatGPT, during the preparation of this manuscript to assist with language refinement, readability, and clarity. All AI-assisted content was critically reviewed, fact-checked, and curated by the authors. The authors take full responsibility for the accuracy, integrity, and originality of the final manuscript.

## Data Availability

The complete methodology, reproducibility data, statistical results, and validation outputs are available in the RCT-Reviewer Validation repository on GitHub (https://github.com/RCT-Reviewer/Validation) and archived on Zenodo (https://doi.org/10.5281/zenodo.22260983). The modernized tool is available at (https://github.com/aurumz-rgb/RCT-Reviewer). The Tian et al. reference dataset is publicly available on OSF (https://osf.io/k6w9q). Due to publisher licensing restrictions, the exact PDF corpora are not publicly redistributed but are available upon reasonable request via restricted Zenodo access: RCT-Reviewer Corpus (1,000-PDF Tier D corpus, https://doi.org/10.5281/zenodo.22260255) and RCT-Reviewer Corpus 2 (Tier E open-access trial subset, https://doi.org/10.5281/zenodo.22286384).

## Acknowledgements

The authors extend sincere gratitude to Iain J. Marshall, Joël Kuiper, Edward Banner, and Byron C. Wallace for their foundational work in biomedical NLP and for releasing the original RobotReviewer project as open-source.

